# Nanopore metagenomic sequencing of influenza virus directly from respiratory samples: diagnosis, drug resistance and nosocomial transmission

**DOI:** 10.1101/2020.04.21.20073072

**Authors:** Yifei Xu, Kuiama Lewandowski, Louise O Downs, James Kavanagh, Thomas Hender, Sheila Lumley, Katie Jeffery, Dona Foster, Nicholas D Sanderson, Ali Vaughan, Marcus Morgan, Richard Vipond, Miles Carroll, Timothy Peto, Derrick Crook, A Sarah Walker, Philippa C Matthews, Steven T Pullan

**Author notes:** **Corresponding author:** Yifei Xu, **Corresponding author email:**. YX and KL contributed equally, PCM and STP contributed equally.

## Abstract

**Background:** Influenza virus presents a significant challenge to public health by causing seasonal epidemics and occasional pandemics. Nanopore metagenomic sequencing has the potential to be deployed for near-patient testing, providing rapid diagnosis of infection, rationalising antimicrobial therapy, and supporting interventions for infection control. This study aimed to evaluate the applicability of this sequencing approach as a routine laboratory test for influenza in clinical settings.

**Methods:** We conducted Nanopore metagenomic sequencing for 180 respiratory samples from a UK hospital during the 2018/19 influenza season, and compared results to routine molecular diagnostic testing. We investigated drug resistance, genetic diversity, and nosocomial transmission using influenza sequence data.

**Results:** Metagenomic sequencing was 83% (75/90) sensitive and 93% (84/90) specific for detecting influenza A viruses compared with the diagnostic standard (Cepheid Xpress/BioFire FilmArray Respiratory Panel). We identified a H3N2 genome with the oseltamivir resistant S331R mutation in the NA protein, potentially associated with the emergence of a distinct intra-subtype reassortant. Whole genome phylogeny refuted suspicions of a transmission cluster in the infectious diseases ward, but identified two other clusters that likely reflected nosocomial transmission, associated with a predominant strain circulating in the community. We also detected a range of other potentially pathogenic viruses and bacteria from the metagenome.

**Conclusion:** Nanopore metagenomic sequencing can detect the emergence of novel variants and drug resistance, providing timely insights into antimicrobial stewardship and vaccine design. Generation of full genomes can contribute to the investigation and management of nosocomial outbreaks.

## INTRODUCTION

Influenza A viruses (IAV) are enveloped viruses of the *Orthomyxoviridae* family, with a segmented, ∼13kb RNA genome [1,2]. IAV can cause both seasonal epidemics and occasional pandemics, presenting a significant challenge to public health [3]. Seasonal epidemics are estimated to cause half a million deaths globally each year, primarily among young children and the elderly [4]. Estimates suggest a future pandemic could infect 20% to 40% of the world population and cause over 30 million deaths within six months [5,6]. Tracking and characterization of circulating influenza viruses, in both human and animal populations, is critical to provide early warning of the emergence of novel variants with high virulence and to inform vaccine design.

Direct-from-sample metagenomic sequencing can potentially identify all viral and bacterial pathogens within an individual clinical sample. The genomic information generated can comprehensively characterize the pathogens and enable investigation of epidemiology and transmission. Oxford Nanopore Technology (ONT) is a third generation sequencing technology that can generate long-read data in real-time, which has been successfully applied in the real-time surveillance of Ebola, Zika, and Lassa outbreaks [7–9]. ONT Metagenomic sequencing has the potential to be deployed for near-patient testing, providing rapid and accurate diagnosis of infection [10], informing antimicrobial therapy [11–13], and supporting interventions for infection prevention and control [14]. We have recently demonstrated proof-of-principle for a direct-from-sample Nanopore metagenomic sequencing protocol for influenza viruses with 83% sensitivity and 100% specificity compared to routine clinical diagnostic testing [15].

Here we describe Nanopore metagenomic sequencing directly from clinical respiratory samples at a UK hospital during the 2018/19 influenza season, evaluating the applicability of this approach in a routine laboratory as a test for influenza, and investigating where further optimisation is still required before the assay can be deployed in clinical practice. We assessed the performance of this experimental protocol head-to-head with routine clinical laboratory tests, and used the influenza sequence data to investigate drug resistance, genetic diversity, and nosocomial transmission events, demonstrating the diverse benefits that can be gained from a metagenomic approach to diagnostics.

## MATERIALS AND METHODS

### Sample collection from clinical diagnostic laboratory

Residual material was collected from anonymised throat swabs, nasal swabs, and nasopharyngeal aspirates that had been submitted to the clinical diagnostic laboratory at the Oxford University Hospitals NHS Foundation Trust during the 2018/19 influenza season.

Prior to metagenomic sequencing, samples had been tested in the diagnostic laboratory based on a standard operating protocol using either Xpert Xpress Flu/RSV assay (Cepheid, Sunnyvale, CA, USA, that detects influenza A/B and respiratory syncytial virus), or BioFire® FilmArray® Respiratory Panel 2 assay (BioFire Diagnostics, Salt Lake City, UT, USA, that detects a panel of viral and bacterial respiratory pathogens). Xpert reports a quantitative diagnostic result (Ct value) for the detected pathogen, while BioFire® RP2 reports a binary result (pathogen detected or not detected). The diagnostic laboratory routinely applies the BioFire® RP2 assay to samples from a defined subgroup of patients most at risk of severe, complicated, or atypical disease (those with immunocompromise, under the care of infection and respiratory teams, or admitted to critical care units).

### Sample selection for Nanopore metagenomic sequencing

The first laboratory diagnosis of influenza in our hospital laboratory in the 2018/19 season was made on 30th October 2018, and our sample collection ran until 5th February 2019. During this period, 1,789 respiratory samples were submitted to the diagnostic laboratory and tested by Xpress Flu/RSV assay, of which 213 were positive for IAV (11.9%); 752 samples were tested by BioFire® FilmArray® Respiratory Panel 2 assay, of which 27 were positive for IAV (3.5%).

90 samples positive for influenza (based on results from Xpress Flu/RSV) and 90 samples negative for influenza (based on results from BioFire® RP2 assay) were selected for Nanopore metagenomic sequencing as follows (Figure 1 and Table S1):

**Figure 1.**
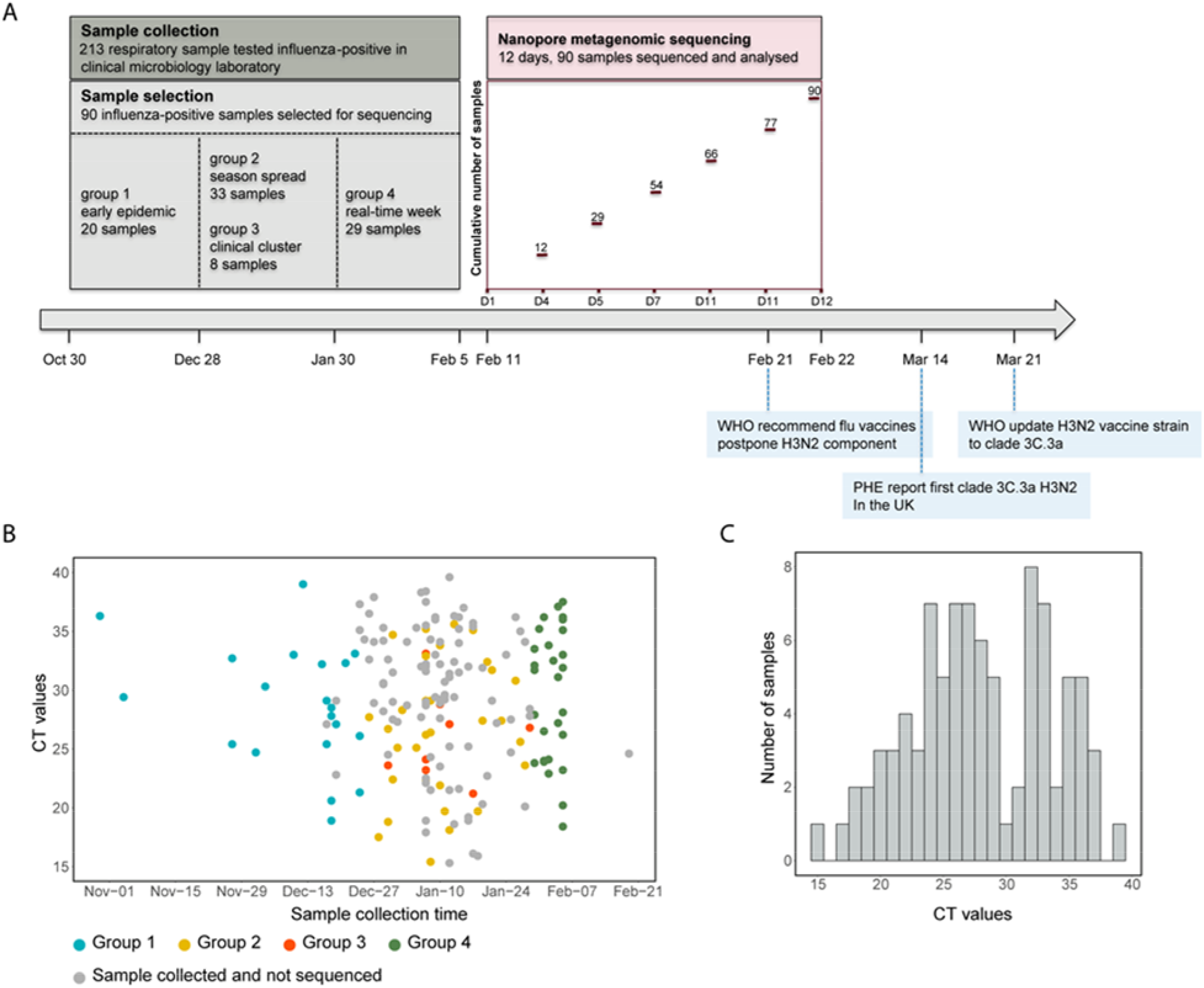
Overview of Nanopore metagenomic sequencing of respiratory samples submitted to the clinical diagnostic laboratory at the Oxford University Hospitals NHS Foundation Trust during the 2018/19 influenza season. A) Timeline of sample collection, sample selection, and Nanopore sequencing. B) Distribution of 90 influenza-positive samples selected for sequencing amongst total influenza-positive samples collected. C) Histogram of Ct values of 90 influenza-positive samples selected for sequencing (Ct value range from 15-39; (mean 28)). Ct values were derived from Xpert Xpress Flu/RSV assay (Cepheid) in the clinical diagnostic laboratory.

1. **1. Influenza-positive samples** (Figure 1A and 1B):
  a. Group 1 (n=20): the first 20 positive samples of the influenza season, from 30th October - 24th December 2018.
  b. Group 2 (n=33): randomly selected samples from the intervening period between Group 1 and 4.
  c. Group 3 (n=8): samples from a putative transmission cluster on the infectious diseases ward diagnosed between 29th December 2018 and 29th January 2019.
  d. Group 4 (n=29): all influenza positive samples from the week beginning 30th January 2019 immediately prior to the onset of sequencing in this study, to represent consecutive samples from a single week at the peak of the influenza season.
2. **Influenza-negative samples:**
  a. 55 samples positive for one of the following viruses: coronavirus (n=10), rhino/enterovirus (n=20), human metapneumovirus (HMPV) (n=5), parainfluenza (PIV) (n=10), and respiratory syncytial virus (RSV) (n=10). Among them, one was positive for both RSV and HMPV, another was positive for PIV, HMPV, and Adenovirus.
  b. 35 samples negative for all pathogens tested by the BioFire® panel.

Methods for sample processing (sequence independent single primer amplification as described in [15]), Nanopore sequencing, and genomic and phylogenetic analyses are described in supplementary material.

## RESULTS

### Nanopore sequencing of influenza directly from respiratory samples

For the Nanopore metagenomic sequencing, the sample processing and library preparation time in our protocol was eight hours, the sequencing time was 48 hours, and thus total turnaround time for each sample was <72 hours. With a team of three members, we prepared the sequencing libraries for 180 samples within 9 days and completed the sequencing runs for the 90 influenza-positive samples within 12 days of commencing sequencing (Figure 1A).

Nanopore sequencing generated between 4.9×10^3^ and 4.1×10^6^ (mean 4.3×10^5^) total reads per sample (Table S1). We retrieved Hazara virus reads (spiked as an internal control at 10^4^ genome copies/ml) from 147/180 (82%) samples. The 33 samples in which Hazara virus reads were not identified were all influenza negative and had comparatively low total cDNA concentrations following amplification. Therefore, we repeated sequencing of the 18/33 samples that had sufficient remaining material with the addition of linear polyacrylamide as a carrier, which produced Hazara virus reads in 16/18 samples. Taken together, we therefore retrieved Hazard internal control in 163/180 (91%) samples (15 were not possible to re-test with carrier).

### Identification, subtyping, and recovery of IAV genomes

The Xpert Xpress Flu/RSV assay (Cepheid, Sunnyvale, CA, USA) reported Ct values ranging from 15.4 to 39.0 (mean 28.0) in the 90 influenza-positive samples, distributed across the flu season (Figure 1B and 1C). We identified IAV reads in 75/90 influenza-positive samples (sensitivity 83%), ranging from 1 to 171,733 reads (Figure 2A). IAV reads were present in all 58 samples with Ct ≤31, and up to a maximum Ct value of 36.3 (sample 48, 12 IAV reads). There was a strong correlation between Ct value and both IAV read numbers (R^2^ =0.43, p<0.0001; Figure 2A) and the ratio of IAV:Hazara virus reads (R^2^ =0.54, p<0.0001; Figure 2B). The remaining 15 influenza-positive samples for which IAV reads were not generated by Nanopore sequencing had lower viral titres, reflected by higher Ct values (range 31.7-39.0). IAV reads were not present in 84/90 influenza-negative samples (specificity 93%).

**Figure 2.**
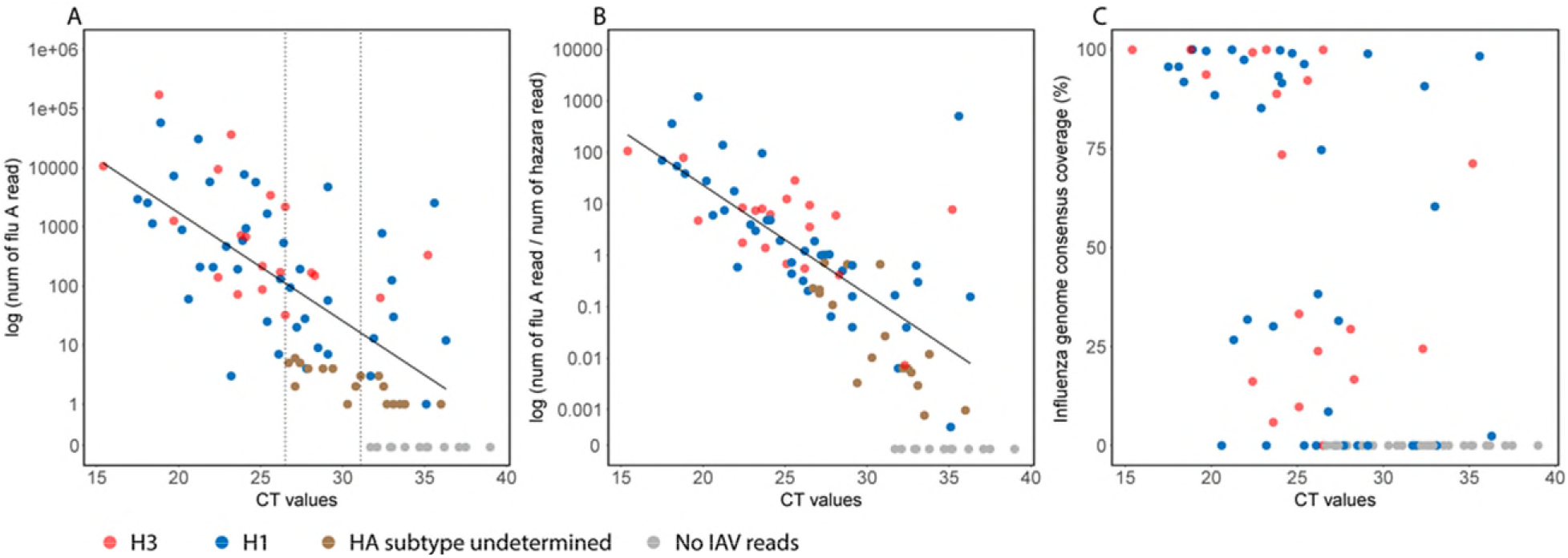
Identification, subtyping, and recovery of IAV genomes by Nanopore metagenomic sequencing of 90 respiratory samples that tested influenza-positive in a UK clinical diagnostic laboratory during the 2018/19 influenza season. Ct values were derived from the routine diagnostic test (Cepheid Xpert Xpress Flu/RSV assay. A) Number of IAV reads generated by Nanopore sequencing against Ct value; R^2^ =0.43, p<0.0001. IAV reads were present in all samples with Ct ≤31 (right dashed line), and HA subtype was determined for all samples with Ct ≤27 (left dashed line). B) Ratio of the number of IAV:Hazara virus reads against Ct value; R^2^ =0.54, p<0.0001. Hazara virus was spiked as an internal control at 10^4^ genome copies/ml. C) Coverage of the IAV consensus sequence against Ct value.

Among the 75 samples for which we generated IAV reads, we could determine the HA subtype of 59/75 (79%) samples; 40 were H1 and 19 were H3 (designated as blue vs red dots in Figure 2). We could determine HA subtype for all samples with Ct ≤27, and up to a maximum of Ct 36.3 (sample 48) (Figure 2A). We retrieved 28/75 (37%) consensus sequences with genome coverage ≥70%, among which 18 were H1 and 10 were H3 subtype (Figure 2C). The genome coverage for samples with Ct value between 20 and 25 showed substantial variation, which was not associated with any sample attributes that we were able to measure, including sample type, or percentage of human or bacterial reads (data not shown).

### Identification of drug-resistant mutations

From consensus sequences covering drug-resistant positions, we identified the S31N amino acid mutation in the M2 protein in 20/20 H1N1 and 11/11 H3N2 sequences, which is known to be widespread, conferring reduced inhibition by amantadine [16]. 1/13 H3N2 sequences (sample 5) carried the S331R amino acid mutation in the NA protein, which has been reported to confer reduced inhibition by oseltamivir [17]. Analysing mapping data for sample 5, 51/53 (96%) reads carried the S331R mutation. Other drug resistance mutations, such as H275Y in the NA protein associated with oseltamivir resistance [18], were not present in our dataset.

### Identification of H3N2 reassortant IAV

The majority of our H3 sequences were clustered within clade 3C.2a1b, with one sequence in clade 3C.3a (Figure 3A). Comparison of the H3 and N2 phylogenies showed that HA and NA segments of each individual sample were clustered within the same clade, except sample 5 had a distinct genotype with the H3 segment clustered within clade 3C.2a1b and the NA segment within clade 3C.2a2 (denoted subsequently as ‘R-genotype’), suggesting intra-subtype reassortment (Figure 3A and 3B).

**Figure 3.**
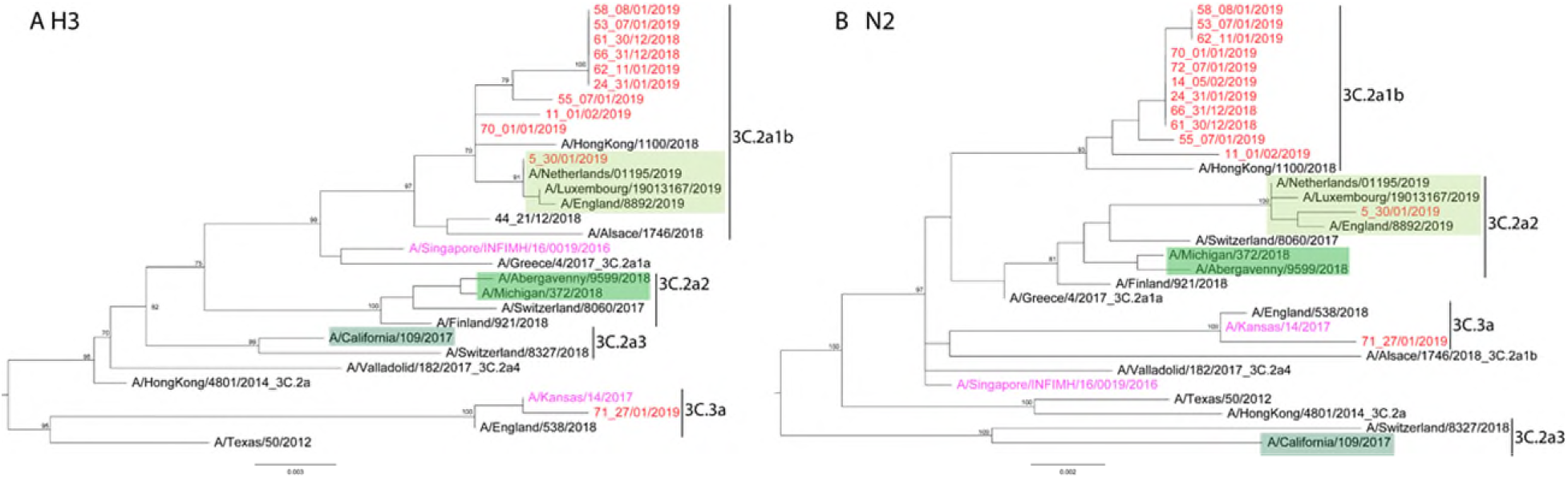
Maximum likelihood phylogenies of H3N2 IAV sequences recovered from respiratory samples collected from a UK hospital cohort during the 2018/19 influenza season. A) H3 segment; B) N2 segment. Sequences recovered from this study are marked in red, vaccine seed strains recommended by WHO for the northern hemisphere 2018/19 and 2019/20 influenza season are in purple, and reference sequences representing major genetic clades of seasonal H3N2 are in black. Genetic clades are indicated on the right of the tree. Green boxes in different shades indicate sequences that carry the S331R mutation in the NA segment: the light green box represent sequences from the 2018/19 season, and two darker green boxes represent sequences from the 2017/18 season.

Interestingly, the S331R mutation occurred in the same sample (sample 5), motivating us to further investigate the prevalence of this mutation in seasonal IAV using all published H3N2 sequences from the last two influenza seasons (2017/18 and 2018/19). In the 2017/18 dataset, 13/7129 (0.2%) sequences carried the S331R mutation, with HA and NA segments from clade 3C.2a2 or 3C.2a3. In 2018/19, the proportion of sequences with the S331R mutation increased to 139/9274 (1.5%), and all belonged to the R-genotype. These results suggest a potential association between the increase in prevalence of the S331R mutation and the emergence of this distinct R-genotype.

### Nosocomial transmission of H3N2 IAV

We included a putative clinical cluster of eight influenza-positive samples (group 3) collected from patients on the infectious diseases ward over a 30 day period, aiming to investigate potential nosocomial transmission events (Figure 4A). We could determine the HA subtype of six samples, three being H3 and three H1. Among these, two H3N2 (samples 53 and 55) and one H1N1 consensus sequences had >70% full genome coverage. A minimum spanning tree (MST) of our H3N2 sequences showed that samples 53 and 55 differed by 25 SNPs (Figure 4B), despite being collected on the same day from patients on the ward. These results refuted the suspicion that these eight samples from the infectious diseases ward were all associated with a single nosocomial transmission cluster, and suggested that some, if not all, of the patients have acquired influenza infection independently.

**Figure 4.**
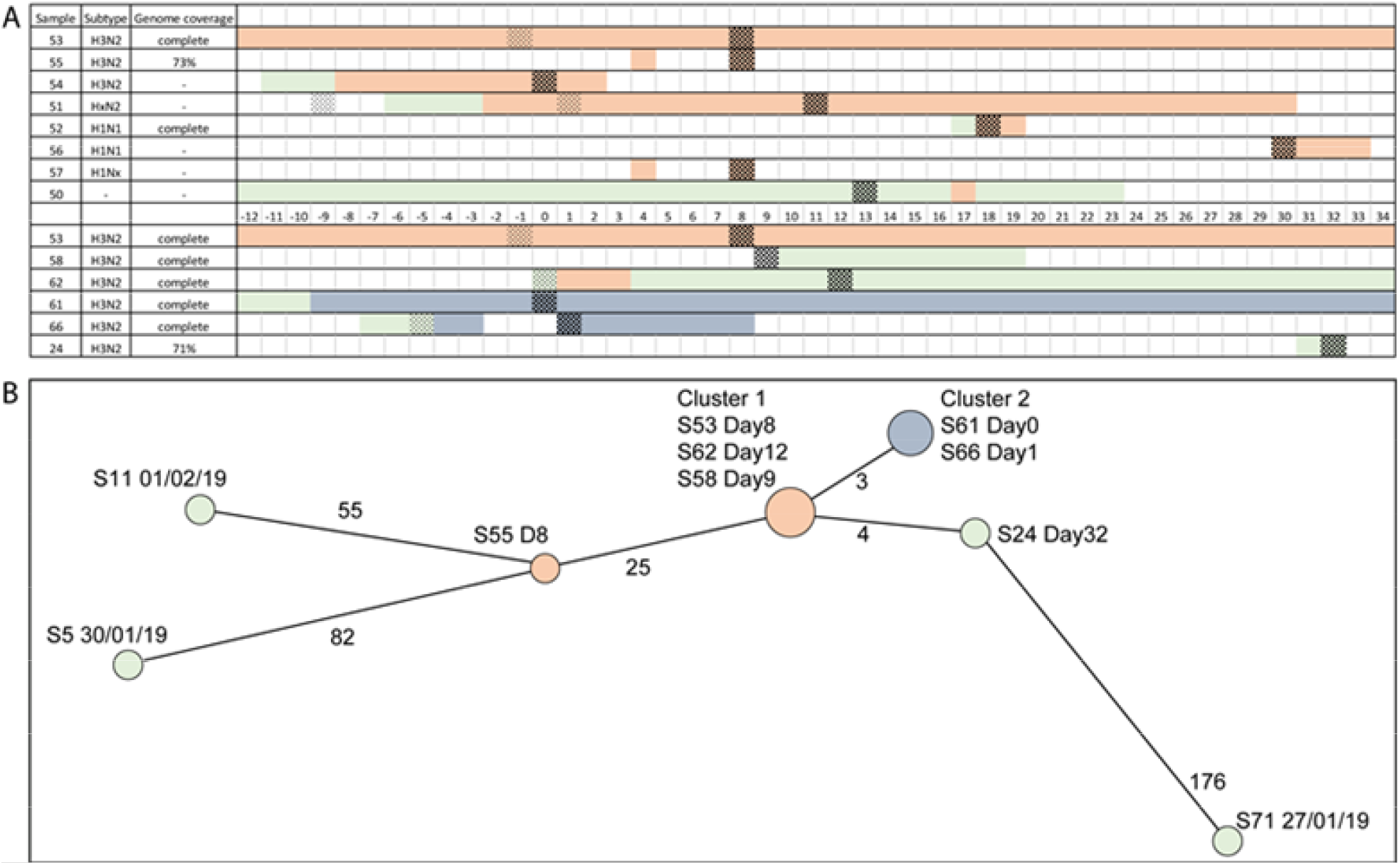
Investigation of nosocomial transmission of H3N2 IAV in a UK hospital during the 2018/19 influenza season. A) Timeline of patients relevant to nosocomial transmission. Each row represents one patient. Timeline indicated in days in the middle divides the plot into two parts: the top part represents eight patients from a putative transmission cluster on the infectious diseases ward, and the bottom part represents six patients with potential nosocomial transmission based on Nanopore sequencing results. Patient’s attendance/admission (in A) and sample (in B) from the infectious diseases ward are in orange, chest ward are in blue, elsewhere in the hospital are in light green. Dark cross hatching box indicates collection of a sample that was tested positive for influenza, and light cross hatching box indicates a sample that tested negative for influenza. B) Minimum spanning tree of H3N2 genomes (coverage >70%). The tree was built on the basis of single nucleotide variant distances between consensus sequences. Distance between each pair of sequence is denoted by number adjacent to the branch.

The H3N2 MST (Figure 4B) also demonstrated:

- Three sequences were identical (cluster 1), from one patient on the infectious diseases ward (sample 53), one who had been recently on the infectious diseases ward and then under the care of emergency assessment unit (EAU) (sample 62), and one who had been on the EAU for a couple of days and then in the complex medicine unit until discharged (sample 58).
- Two identical sequences (cluster 2) differed from cluster 1 by 3 SNPs, and were from patients on the respiratory ward, taken two days apart.
- One sequence (sample 24) differed from cluster 1 by 4 SNPs, and was from an acutely admitted patient in the EAU three weeks later.
- The remaining four sequences, including sample 55 from the refuted cluster and three from patients elsewhere in the hospital, were separated from cluster 1, cluster 2, and each other by ≥25 SNPs.

These results suggested that cluster 1 patients on the infectious diseases ward and cluster 2 patients on the chest ward likely reflected nosocomial transmission. There was no clear link between cluster 1 patients, cluster 2 patients, and the acutely admitted EAU patient (sample 24). One patient in cluster 1 (sample 58) and this EAU patient were positive for influenza on the first day of their admission to the hospital, suggesting these samples may be associated with a predominant strain circulating in the community.

### Independent introductions of pH1N1 IAVs

Phylogenetic analysis of the H1 segment showed that our sequences clustered within clade 6B.1 (Figure S2A). At the full genome level, we found no evidence of phylogenetic clustering of pH1N1 IAVs recovered from our hospital, suggesting these represent independent introductions. Rather, our pH1N1 genomes were closely related to other genomes recovered during the UK 2018/19 season (Figure S2B). 12 of our pH1N1 genomes had their most closely related sequence within <15 SNPs, 11 of these most closely related sequences were from the UK.

### Pilot study of testing for five other respiratory viruses

Among the 90 influenza-negative respiratory samples we sequenced, 55 had tested positive for another virus in the clinical diagnostic laboratory. From this small dataset, our metagenomic sequencing dataset was >80% sensitive for HMPV, RSV, and PIV, but only 30% sensitive for Coronavirus and Enterovirus; specificity was high at >94% for all five viruses (Table 1).

**Table 1.** Summary of results for five respiratory viruses derived from Nanopore sequencing data of 90 respiratory samples collected from a UK hospital cohort. Samples were tested in the clinical diagnostic laboratory using BioFire® FilmArray® Respiratory Panel assay (BioFire Diagnostics, Salt Lake City, UT, USA) for a panel of respiratory pathogens. True and false positive and negative results pertain to results of Nanopore sequencing.

| **Virus** | **Number positive**<br>**based on Biofire**<br>**testing in clinical lab** | **True**<br>**positive** | **False**<br>**negative** | **Sensitivity**<br>**%** | **True**<br>**negative** | **False**<br>**positive** | **Specificity**<br>**%** |
| --- | --- | --- | --- | --- | --- | --- | --- |
| Human Metapneumovirus | 5 | 4 | 1 | 80 | 83 | 2 | 98 |
| Respiratory syncytial virus | 11 | 9 | 2 | 82 | 75 | 4 | 95 |
| Parainfluenza | 11 | 9 | 2 | 82 | 77 | 2 | 97 |
| Coronavirus | 10 | 3 | 7 | 30 | 79 | 1 | 99 |
| Enterovirus | 20 | 6 | 14 | 30 | 68 | 2 | 97 |

### Identification of organisms not tested for in the clinical laboratory

In five influenza-positive samples for which IAV reads were generated by sequencing, we also retrieved reads for other viruses, including human coronavirus HKU1 (sample 17, 996 reads covering the complete genome; sample 14, 76 reads), human parainfluenza virus 3 (sample 40, 3 reads), rhinovirus A (sample 10, 1 read), and human astrovirus (sample 19, 1 read) (Table S1). For the 90 influenza-negative samples, sequencing data did not show reads likely to represent viral pathogens other than those already identified by BioFire.

From our complete collection of 180 samples, we identified reads from five bacterial species, *Streptococcus pneumoniae* (n=37 samples), *Pseudomonas aeruginosa* (n=5), *Moraxella catarrhalis* (n=3), *Staphylococcus aureus* (n=1), and *Haemophilus influenzae* (n=1) (Table S1). While these organisms may represent agents of respiratory infection, they can also be commensal or colonising flora. In the absence of detailed clinical metadata, we were unable to explore their likely contribution to pathology.

## DISCUSSION

### Turnaround time for metagenomic sequencing

In this study, we conducted Nanopore metagenomic sequencing of IAV directly from clinical respiratory samples at a UK hospital during the 2018/19 influenza season, reporting a head-to-head comparison with routine clinical diagnostic tests. The total turnaround time for metagenomic sequencing of each sample was <72 hours. We processed 180 clinical samples and generated sequencing data for 90 clinical samples over a 12 day period. While the turnaround time is still slower than other laboratory diagnostic tests, and the approach requires an investment in labour and sequencing/analysis time, there is potential to reduce this further, through simplification of the wet laboratory protocol and implementation of real-time bioinformatic analysis of reads as they are generated.

Timeliness is crucial for the deployment of international vaccine strategies. Each February, WHO determines influenza vaccines for use in the following northern hemisphere influenza season. However, in 2019, WHO postponed the vaccine update until late March to include a clade 3C.3a H3N2 strain (Figure 1), due to the substantial increase of 3C.3a viruses in several regions since November 2018 associated with low vaccine effectiveness (5%) [19]. This one-month delay raised concerns about the timeliness of vaccine manufacturing and distribution for the upcoming influenza season. Within our cohort, a clade 3C.3a H3N2 sample was collected on 27th January 2019, and if we had conducted rapid-turn-around sequencing as a routine assay then the complete genome sequence could be available in <72 hours, while the first strain from this clade in the country was not officially reported by Public Health England until 14th March. This timeline illustrates how routine laboratory sequencing would allow earlier genetic characterization, providing translational advantages in influenza surveillance, monitoring change in the proportion of genetically diverse strains, and contributing to timely insights into seansonal epidemiology vaccine design.

### Sensitivity of Nanopore metagenomic sequencing

Our sequencing data showed 83% sensitivity for IAV compared with existing laboratory diagnostic tests, consistent with our previous study with a smaller dataset [15]. Further optimization is needed to improve the sensitivity of our protocol for clinical samples with lower viral titres (Ct values >30). Potential methods include depletion of host and bacterial RNA to reduce the amount of non-target nucleic acid present, and enrichment of the target via probes or primer amplification. Our data show that addition of a carrier can improve the detection of internal spiked control in samples with low total cDNA, which is likely due to the improved purification and reduced degradation of lower concentration RNA, thus we intend to incorporate this approach as a routine part of the protocol in future.

### Drug resistance

The S331R NA mutation in H3N2 IAV has been associated with reduced susceptibility to oseltamivir since the 2013/14 influenza season [17,20,21]. Among 1,039 H3N2 IAVs tested globally during the 2018/19 season, one strain from South Korea showed reduced susceptibility to oseltamivir due to this mutation [22]. Our analysis demonstrates that IAVs carrying this mutation from the 2018/19 season belong to a distinct genotype generated through intra-subtype reassortment between clades 3C.2a1b and 3C.2a2. A previous study reported a similar observation that the emergence and rapid global spread of adamantane resistant H3N2 IAVs (conferred by a S31N mutation in the M2 protein) was associated with a single genotype generated through intra-subtype reassortment [23,24]. S31N now occurs in almost all circulating IAV globally, causing the cessation of use of adamantane to treat influenza [16]. The genesis, prevalence, distribution and clinical impact of the S331R mutation merits additional study to evaluate potential implications for the clinical usefulness of oseltamivir, which is widely used as a first-line agent when treatment is indicated [20].

### Mapping outbreaks and transmission

Whole genome sequencing can provide high resolution characterization of the spatiotemporal spread of viral outbreaks [7,8]. Previous studies have used targeted enrichment combined with next generation sequencing to investigate nosocomial transmission of influenza [14,25], and our study demonstrates the application of Nanopore metagenomic sequencing for this purpose. Our sequencing data allow us to refute the suspicion of a single transmission cluster on the infectious diseases ward, although the small number of whole genomes generated limits the extent to which we could draw conclusions about transmission among this specific patient group. Furthermore, our dataset reveals two clinical clusters that likely represent nosocomial transmission on the infectious diseases ward and the chest ward, showing proof of concept that Nanopore metagenomic sequencing can identify nosocomial transmission with the potential to inform infection prevention and control practices.

### Detection of organisms other than IAV

Based on a small exploratory dataset, our protocol shows >80% sensitivity for the detection of human metapneumovirus, parainfluenza, and respiratory syncytial virus compared to routine clinical diagnostic testing. The lower sensitivity for enterovirus and coronavirus could be due to low viral titres in these samples, although we are not able to confirm this as the BioFire® RP2 assay is a non-quantitative test. Another possibility is that the SISPA method is less sensitive for certain viruses [26]. Moreover, no influenza B virus reads are present in our 90 influenza-positive samples, congruent with the global low level of influenza B virus during the 2018/19 season. Further work is needed to determine the limits of detection and optimize the laboratory and bioinformatic protocol to improve the sensitivity for a wider range of potential pathogenic organisms.

### Caveats and limitations

This study included a limited cohort, with samples stratified by clinical diagnostic results, collection time, and the observation of a putative clinical cluster. We were not able to systematically sequence all influenza-positive samples from the clinical diagnostic laboratory due to limited manpower and laboratory resources. Generalisability is limited by this sampling approach, as well as by other confounding influences which we were unable to control, including both laboratory and clinical influences (e.g. diverse sample types, sample exposure to freeze/thawing, underlying immunocompromise, symptom duration prior to sample collection).

While metagenomic data holds the promise for simultaneous detection of all pathogens from an individual clinical sample, it poses general challenges to analyze and distinguish between pathogens, commensal flora and potential contaminants. Accurate interpretation is based upon the clinical context of the patient, type and quality of the sample, the absolute and relative abundance of the organism in the metagenome, genome coverage and mapping depth, and the occurrence of the organism in samples on the same run (if multiplexed) and historical runs in the same laboratory. Expert case-by-case appraisal is currently required if the data are to be used for clinical decision-making.

## Conclusions

In summary, we demonstrate the feasibility of applying Nanopore sequencing in clinical settings to simultaneously detect influenza and other respiratory viruses, identify drug resistance mutations, characterize genetic diversity, and investigate potential nosocomial transmission events. While work is still needed to refine and streamline the sequencing protocol and bioinformatic analysis, Nanopore metagenomic sequencing has the potential to become an applicable point-of-care testing for infectious diseases in clinical settings.

## Data Availability

Data availability
Following removal of human reads, our sequencing data have been uploaded to the European Bioinformatics Institute https://www.ebi.ac.uk/, project reference PRJEB…..

## Ethical statement

The study of anonymised discarded clinical samples was approved by the London - Queen Square Research Ethics Committee (17/LO/1420).

## Funding statement

The study was funded by the NIHR Oxford Biomedical Research Centre. Computation used the Oxford Biomedical Research Computing (BMRC) facility, a joint development between the Wellcome Centre for Human Genetics and the Big Data Institute supported by Health Data Research UK and the NIHR Oxford Biomedical Research Centre. The views expressed in this publication are those of the authors and not necessarily those of the NHS, the National Institute for Health Research, the Department of Health or Public Health England. PCM is funded by the Wellcome Trust (grant ref 110110) and holds an NIHR senior fellowship award. DWC, TEAP and ASW are NIHR Senior Investigators.

### Acknowledgements

We gratefully acknowledge input from, and support of, all the members of the microbiology laboratory team and the infection prevention and control team at Oxford University Hospitals NHS Foundation Trust.

## Data availability

Following removal of human reads, our sequencing data have been uploaded to the European Bioinformatics Institute https://www.ebi.ac.uk/, project reference PRJEB…..

